# Yet another lockdown? A large-scale study on people’s unwillingness to be confined during the first 5 months of the COVID-19 pandemic in Spain

**DOI:** 10.1101/2021.05.08.21256792

**Authors:** Marina Martinez-Garcia, Alejandro Rabasa, Xavier Barber, Kristina Polotskaya, Kristof Roomp, Nuria Oliver

## Abstract

Population confinements have been one of the most widely adopted non-pharmaceutical interventions (NPIs) implemented by governments across the globe to help contain the spread of the SARS-CoV-2 virus. While confinement measures have been proven to be effective to reduce the number of infections, they entail significant economic and social costs. Thus, different policy makers and social groups have exhibited varying levels of acceptance of this type of measures. In this context, understanding the factors that determine the willingness of individuals to be confined during a pandemic is of paramount importance, particularly, to policy and decision-makers. In this paper, we study the factors that influence the unwillingness to be confined during the COVID-19 pandemic by means of a large-scale, online population survey deployed in Spain. We apply both quantitative (logistic regression) and qualitative (automatic pattern discovery) methods and consider socio-demographic, economic and psychological factors, together with the 14-day cumulative incidence per 100,000 inhabitants. Our analysis of 109,515 answers to the survey covers data spanning over a 5-month time period to shed light on the impact of the passage of time. We find evidence of *pandemic fatigue* as the percentage of those who report an unwillingness to be in confinement increases over time; we identify significant *gender* differences, with women being generally less likely than men to be able to sustain long-term confinement of at least 6 months; we uncover that the *psychological impact* was the most important factor to determine the willingness to be in confinement at the beginning of the pandemic, to be replaced by the *economic impact* as the most important variable towards the end of our period of study. Our results highlight the need to design gender and age specific public policies, to implement psychological and economic support programs and to address the evident pandemic fatigue as the success of potential future confinements will depend on the population’s willingness to comply with them.

## Introduction and Related work

Population confinements have been one of the most widely adopted non-pharmaceutical interventions (NPIs) implemented by governments across the globe to help contain the spread of the SARS-CoV-2 virus^1–4^. While confinement measures have been proven to be effective to reduce the number of infections^3,5–7^, they entail significant economic and social costs. Thus, different policy makers and social groups have exhibited varying levels of acceptance of this type of measures, from the low levels of mask wearing compliance^8^ and public acceptance of COVID-19 lockdown scenarios^9^ in Germany to the highly interventionist Chinese experience^10,11^. In this context, understanding the factors that determine the willingness of individuals to be confined during a pandemic is of paramount importance, particularly, to policy and decision-makers.

In this paper, we study the factors that determine people’s unwillingness to be confined during the first five months of the COVID-19 pandemic in Spain by means of a large-scale, online population survey. We consider socio-demographic, economic and psychological factors, together with reported 14-day cumulative incidence per 100,000 inhabitants. Our analysis covers data spanning over a 5-month time period to shed light on the impact of the passage of time.

Since the COVID-19 pandemic was declared, numerous studies that have analysed the economic, social and psychological impact of NPIs. Moreover, several authors have reported on people’s experiences during the confinements implemented to contain the spread of coronavirus. We summarize next the most relevant pieces of previous work as they relate to ours.

### Economic impact of NPIs

Regarding the economic impact of non-pharmaceutical interventions, several works have assessed the cost of different types of NPIs in the context of the current coronavirus pandemic. Abiad *et al*. simulate different scenarios of the evolution of the pandemic in order to estimate the economic impact^12^. Fernandes *et al*. maintain the same central idea of studying different scenarios and highlight how different this crisis is (due to the pandemic) compared to previous ones^13^. Possible supply chain scenarios are analyzed in^14^, highlighting that the timing of the closing and opening of the facilities at different stages might become a major factor in determining the impact of epidemic outbreaks on the supply chain performance. Finally, the work by Chen and colleagues^15^ highlights the impact that socio-psychological variables have on household consumption patterns and their ability and willingness to assume higher economic expenses.

### Social impact of NPIs

Beyond the economic dimension, the pandemic has also significantly impacted people’s social lives. Clemens *et al*. asses the impact that social distance and confinement measures have on people’s relationships^16^. Jiao and colleagues^17^ establish guidelines for children and adolescents to mitigate the effect of social isolation during the pandemic. Social vulnerability is pointed out as the main risk of the pandemic by^18^, recommending improvements of mental, social and spiritual resilience in the population. Mukhtar reviews past outbreaks to understand the impact of adverse effects on psychological health, psychological crisis intervention, and mental health management plans^19^. He concludes that individuals (and societies in general) must deliberately cultivate resilience and develop strategies to cope with adversity, in the case of the current SARS-CoV-2 pandemic caused by confinement.

### Psychological and behavioural impact of NPIs

Several studies have analyzed the psychological impact of the confinement measures via online surveys and/or smartphone apps. Gao *et al*.^20^ report a significant increase in media usage during the first wave of the SARS-CoV-2 pandemic in China from a sample of 4,872 individuals collected over a 3-day period (from the 31st of January to the 2nd of February, 2020). Their main findings show that there is a high prevalence of mental health problems, positively associated with frequent exposure to social media during the COVID-19 outbreak. As a result of this finding, the authors suggest that governments should pay more attention to the population’s mental health among the general population while fighting the coronavirus pandemic. Hawryluch et al.^21^ describe the results of analyzing data from 129 individuals collected via a long 152-question survey to capture the psychological impact of the confinement in Canada, during previous SARS-CoV-1 outbreak. They conclude that the prevalence of post-traumatic stress disorder and depressive symptoms grows as the period of confinement increases. A multivariate logistic regression model was used by Wang et al.^10^ to assess the impact of the confinement measures on people’s mental health. The authors observed that the confinement measures adopted by the governments had a negative impact on the mental health of particularly vulnerable groups, including those with previous mental health disorders or chronic physical problems, first-line workers, coronavirus infected or suspected patients and those who had financial problems.

Recent work has also explored the use of technology to provide psychological support to people while in confinement, such as the self-guided activities proposed in^22^, models to manage potential psychological crises and decision support systems to help alleviate psychiatric and psychological disorders caused or exacerbated due to the confinement^23^.

General behavioural patterns and perceptions towards the pandemic and the confinement measures are described in^24^ from a sample of 1,420 individuals collected between the 18th and 24th of March, 2020 in Australia. The study identifies the main difficulties reported by the participants to comply with the measures of physical distancing, namely home sharing, the responsibility of caring for someone outside your home and working from home. Yamada et al. describe the COVIDiSTRESS survey (data collected between April and May 2020) deployed to study the psychological and behavioural responses to the coronavirus pandemic and associated government measures – such as the cancellation of public events and stay-at-home orders implemented in many countries. A global consortium of 39 countries and regions worked together to construct and translate the survey with variables of shared interests and recruited participants in 47 languages^25^. The survey, with 1,304 respondents, covered several areas, including demographic data, physical symptoms in the past 14 days, contact history with COVID-19 in the past 14 days, knowledge and concerns about COVID-19, precautionary measures against COVID-19 in the past 14 days, additional information required with respect to COVID-19, the psychological impact of the COVID-19 outbreak; and mental health status. Finally, Bailon et al.^26^ analyze the longitudinal answers of 1,000 respondents provided over a 2.5 month period in Spain, aiming to understand potential changes in people’s mood during the confinement. Beyond the study on mood variations during confinement, this paper offers interesting descriptive statistics of the socio-demographic variables of the participants, as well as their housing characteristics and physical activity habits.

### Willingness to comply with the confinement measures

Similar to our research, most of the previous work that has studied people’s experience with the COVID-19 pandemic confinement measures has relied on self-reported survey data. Amat et al.^27^ analyze 1,600 answers collected between the 20th and 28th of March, 2020 from Spain and United States. The authors claim that during the coronavirus pandemic individuals have relinquished more rights than during other international crisis scenarios as climate change or international terrorism. Flexibility and the ability to adapt have been found to be key skills to help people cope with confinement situations. Tintori et al.^28^ report such a finding from analysing 140,000 online interviews between the 22nd of March and the 2nd of April, 2020; and Landi et al.^29^ collect data from 944 adults in Italy over 35 days and propose an acceptance and compromise therapy based in the concept of psychological flexibility as a tool to help people during confinement. McBride at al.^30^ propose empirical indicators captured by the Brief Resilience Scale to quantitatively assess people’s acceptance of the confinement measures. They validate the proposed instrument via a study with 2,000 individuals in United States, between the 23rd and the 28th of March, 2020. Other authors, such as^31^, study the impact of conspiracy theories on people’s willingness to comply with the confinement measures. They collect data via an online survey with 2,501 adults in England, quota sampled to match the population for age, gender, income, and region. According to their findings, there is a notable endorsement of conspiracy beliefs about coronavirus in England, and such ideas do not appear to be limited to the fringes. These conspiracy beliefs connect to other forms of mistrust and are associated with less compliance with government guidelines and greater unwillingness to take up future tests and treatment. Finally, Chan et al.^32^ suggest that regions with risk-averse attitudes are more likely to adjust their behavioural activity in response to the declaration of a pandemic even before official government lockdowns.

In view of the reported previous work and given the focus of our research, the main contributions of our paper are four-fold. First, we report the results of analyzing a very large-scale, online population survey with 109,515 answers from Spain collected between April 3rd, 2020 and September 11th, 2020. Second, we apply two different methodologies to predict and shed light on the factors that impact people’s willingness to be confined: logistic regression and pattern-discovery, rule-based models. Third, we consider socio-demographic, economic and psychological factors, together with reported 14-day cumulative incidence per 100,000 inhabitants. Finally, we analyse data covering a 5-month period to understand the impact of the passage of time on people’s unwillingness to be confined.

In the following section, we describe the main results of our analyses. Next, we describe our methodology and discuss the main findings of our work. Finally, we conclude with a summary of our contributions and an outline and future lines of research.

## Results

In this section, we describe the main results of our analyses, starting with the descriptive statistics of the data set, followed by the results of a logistic regression modeling and the patterns that were automatically identified by our pattern identification approach (described in the Methods section).

### Descriptive Statistics

We report the results of analyzing a subset of the answers to the COVID19ImpactSurvey, a very large, anonymous, online citizen survey about COVID-19^33^. Launched on March, 28th 2020 in Spain, the survey has since collected over 600,000 anonymous answers mainly from 4 countries (Spain, Italy, Brazil and Germany). From this very large sample, we analyze the data from Spain for the time period between April, 3rd 2020 and September, 11th 2020.

From an original set of 25 questions, we analyze the answers to the questions included in Table 1 in the Supplementary Information. Given that the data was collected by means of a non-probabilistic sampling method, we weigh the answers such that the distribution of answers per age, gender and geographic province matches the officially reported distributions in the Spanish census data. A similar methodology was implemented in^33^. Table 2 in the Supplementary Information shows the distributions of the age and gender variables before and after weighting the survey answers. The Table also includes the official census data for comparison.

**Table 1.** Independent and dependent variables used in our study.

| Variable/Values | Explanation |
| --- | --- |
| *Willingness to be confined* |  |
| 0 | Willingness to be confined for 0 days |
| 1 | Willingness to be confined for $\geq 6$ months |
| *Phase* |  |
| Workplace closure<br>28th March 2020 | Closure of all non-essential labor |
| Workplace reopening<br>12th April 2020 | Return to work in sectors affected by Workplace closure |
| Walks allowed<br>28th April 2020 | Citizens are allowed to leave their homes to go for walks after domiciliary confinement |
| Phase 0<br>4th May 2020 | Public transport/museums/churches at 33% capacity;<br>Takeaway food open |
| Phase I<br>11th May 2020 | Phase 0 + Reopening of Hotels (except common areas);<br>Restaurant terraces to 50% capacity;<br>Mobility inside the province to travel to second residences |
| Phase II<br>25th May 2020 | Phase I + Reop. non-university classrooms;<br>Restoration/malls(non common areas)/churches at 50%;<br>Reop. beaches; Cultural spaces/ hotels common areas to 33% |
| Phase III<br>8th Jun 2020 | Phase II + Reop. discos without dance floor at 50% capacity;<br>Public transport at 100% capacity;<br>Reop. hotels/malls common areas/cultural spaces at 50% capacity;<br>Mobility inside the autonomous region |
| New Normal (21st Jun 2020 onwards) | Normal activity is resumed; Masks are compulsory in public spaces |
| *Age (Q1)* |  |
| “18-29” | 18 to 29 years old |
| “30-49” | 30 to 49 years old |
| “50-59” | 50 to 59 years old |
| “60+” | 60 or more years old |
| *Gender (Q2)* |  |
| Man | Male respondents |
| Woman | Female respondents |
| *Home (Q5)* |  |
| Apartment | The respondent lives in an apartment |
| Single family | The respondent lives in a single family house |
| *Household (Q6)* |  |
| Young | Every house member is aged $< 60$ years old |
| Elderly | Without any children at home and every house member aged 60+ |
| Multi-generational | With children (aged $< 18$) and members aged 60+ |
| *Economic Impact (Q15)* |  |
| None | No economic impact is reported |
| Mild | I lost my job, I lost part or all of my savings,<br>My business is in danger of bankruptcy<br>My employer is at risk of bankruptcy |
| Severe | I can't afford to buy food, I can't pay my rent or mortgage anymore |
| *Psychological Impact (Q25)* |  |
| None | No psychological impact is reported |
| Mild | Excessive use of technology by adults (tablet, phone, TV),<br>Excessive use of technology by children (tablet, phone, TV) |
| Severe | High levels of anxiety, High levels of stress, High levels of loneliness,<br>High levels of sadness, Loud arguments or fights with other members of the home, Excessive consumption of alcohol,<br>Excessive consumption of drugs (prescription or other) |
| *CumInc* | Average 14-day Cumulative Incidence per 100,000 people in each of the 17 Autonomous Regions in Spain |

**Table 2.**
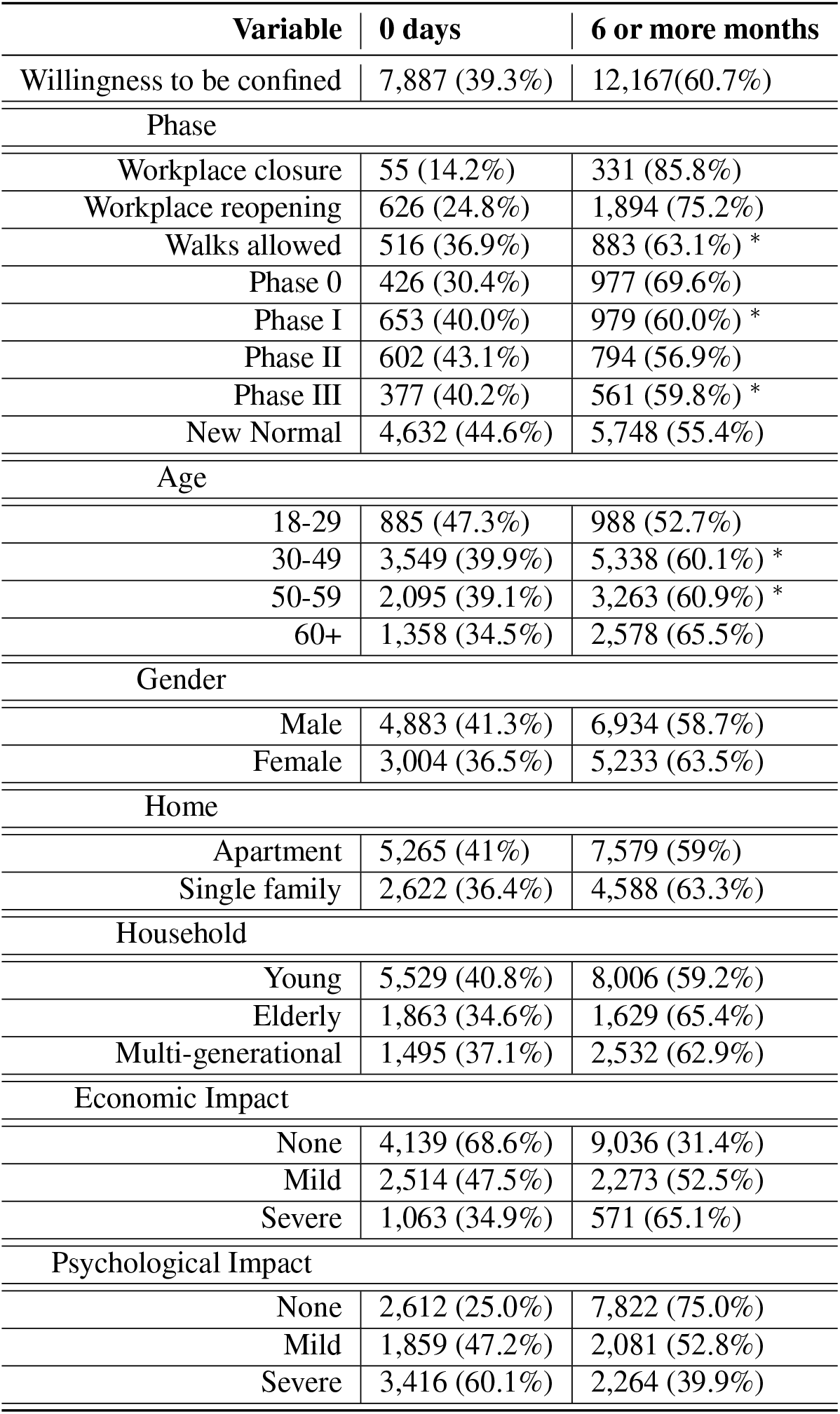
Descriptive statistics of the subset of answers to the survey analyzed in this paper. All the variables pass a proportions test, with p-values *<* 0.001, except for the variables marked with ^*^

Table 1 depicts the dependent and independent variables used in our analysis. We are particularly interested in understanding the differences in demographic factors, impact and perceptions between those who report an opposition to be confined vs those who report a willingness to be confined for at least 6 months. Thus, we model our dependent variable (willingness to be confined) as a binary variable where 0 represents an unwillingness to be confined and 1 represents maximum acceptance towards confinement (willing to be confined for 6 months or more). When we select the answers of those reporting an unwillingness to be confined and those reporting an acceptance to be confined for at least 6 months, we obtain a sample with 20,054 responses corresponding to the time period between April, 3rd and September, 11th 2020.

Table 2 shows the resulting proportions of the variables analysed in the study. As seen on the Table, the data set is unbalanced regarding the target variable: there are 7,887 (39.3%) answers of those reporting an unwillingness to be confined vs 12,167 (60.7%) answers of those who report high willingness towards confinement (≥6 months). In terms of gender, there are 8,237 (41%) and 11,817 (59%) answers by women and men, respectively. We group the age of our participants into four possible age buckets: [18-29], [30-49], [50-59] and 60+, with 9%, 44%, 27% and 20% of the answers, respectively.

To consider the impact of time, we include a discrete variable called *Phase* that refers to the different phases of confinement that were applied in Spain in the spring and summer of 2020, as shown in Figure 5 and described in Table 1. From Table 2, we observe a decrease over time in those reporting a willingness to be confined for a long time period: at the beginning of the pandemic (*Workplace closure* phase), they represent 85.8% of the sample, whereas once the *New normal* phase was declared at the end of June of 2020, this group was only 55.4% of the sample. Thus, we use the Phase variable to segment the sample in 8 different time periods, and we build models both for the entire time period and for each of the confinement phases independently.

We first performed a proportions test to determine if the distribution of answers is significantly different for each of the variables of interest between those unwilling to be confined and those with high levels of acceptance towards confinement. We found statistically significant differences in all the variables (p-value < 0.01) except for the variables marked with ^*^ on Table 2, which correspond to *Walks allowed, Phase I* and *Phase III* and for the age groups *30-49* and *50-59*. Nonetheless, note that all the variables have at least two values with significant differences. Thus, we conclude that all variables are relevant to be included in our models.

The main research questions that we would like to answer through the analysis of this data set are:

1. **RQ1:** Are there differentiating attributes between those who report high vs no willingness to be confined?;
2. **RQ2:** Is it possible to accurately predict an unwillingness to be confined from the independent variables?; and
3. **RQ3:** How did the willingness to be confined evolve during the different phases of the confinement?

### Logistic Regression Model

We obtain a logistic regression model with all the variables described in Table 1 and the interactions shown in Table 3. We refer the reader to the Materials and Methods section for a description of our approach. We compute the McFadden’s pseudo-R squared (0.131), the Cox-Snell residual (0.161) and Nagelkerke’s R squared (0.218) to asses the the goodness of fit of the selected model.

**Table 3.** Significant interactions identified by the logistic regression model.

| Variable | Significant interactions with |
| --- | --- |
| Phase | PsyI, Gender, Household |
| Gender | EcoI, Home, Phase, Household |
| Age | PsyI |
| Home | Gender |
| Household | PsyI, Phase, Gender |
| EcoI | PsyI, Age, Gender |
| PsyI | EcoI, Age, Household, Phase |
| CumInc | – |

As reflected in Table 3, the variables with the most interactions are *Gender* and *PsyI*. The existence of the variable *Phase* in our model, interacting with *PsyI, Gender and Household*, empirically corroborates a temporal dependency of the target variable. Interestingly, the 14-day Cumulative Incidence of COVID-19 cases (*CumInc*) has no significant interaction with any of the variables.

We performed an Odds Ratio (OR) analysis to shed light on the role of each of the variables in the logistic regression model. A variable with an OR > 1 is typically interpreted as a risk factor, *i*.*e*., it is a variable that significantly increases the probability of the target variable to be 1. Conversely, a variable with an OR < 1 decreases the probability for the target variable to be 1. We reverse the coding of the target variable so it represents a *risk*: we code as 1 the answers corresponding to an unwillingness to be confined and as 0 the answers corresponding to an acceptance towards a confinement of at least 6 months.

In our case, the basal categories for the OR analysis are: *Phase: Workplace closure; Gender: Men; Age: 18-29; Home: Apartment; Household: Young; EcoI: None and PsyI: None*, which correspond to the first row of each category in Table 2.

Note that to obtain the OR value in a logistic regression model with second order interactions among some of the variables (*Phase, Gender, Age, Home, Household, EcoI and PsyI* as per Table 3), the OR of the main variable needs to be multiplied by the OR of the interaction variables where the main variable is present.

Figure 1 depicts the OR of the variables with the most important interactions, grouped in four categories to ease their interpretation.

**Figure 1.**
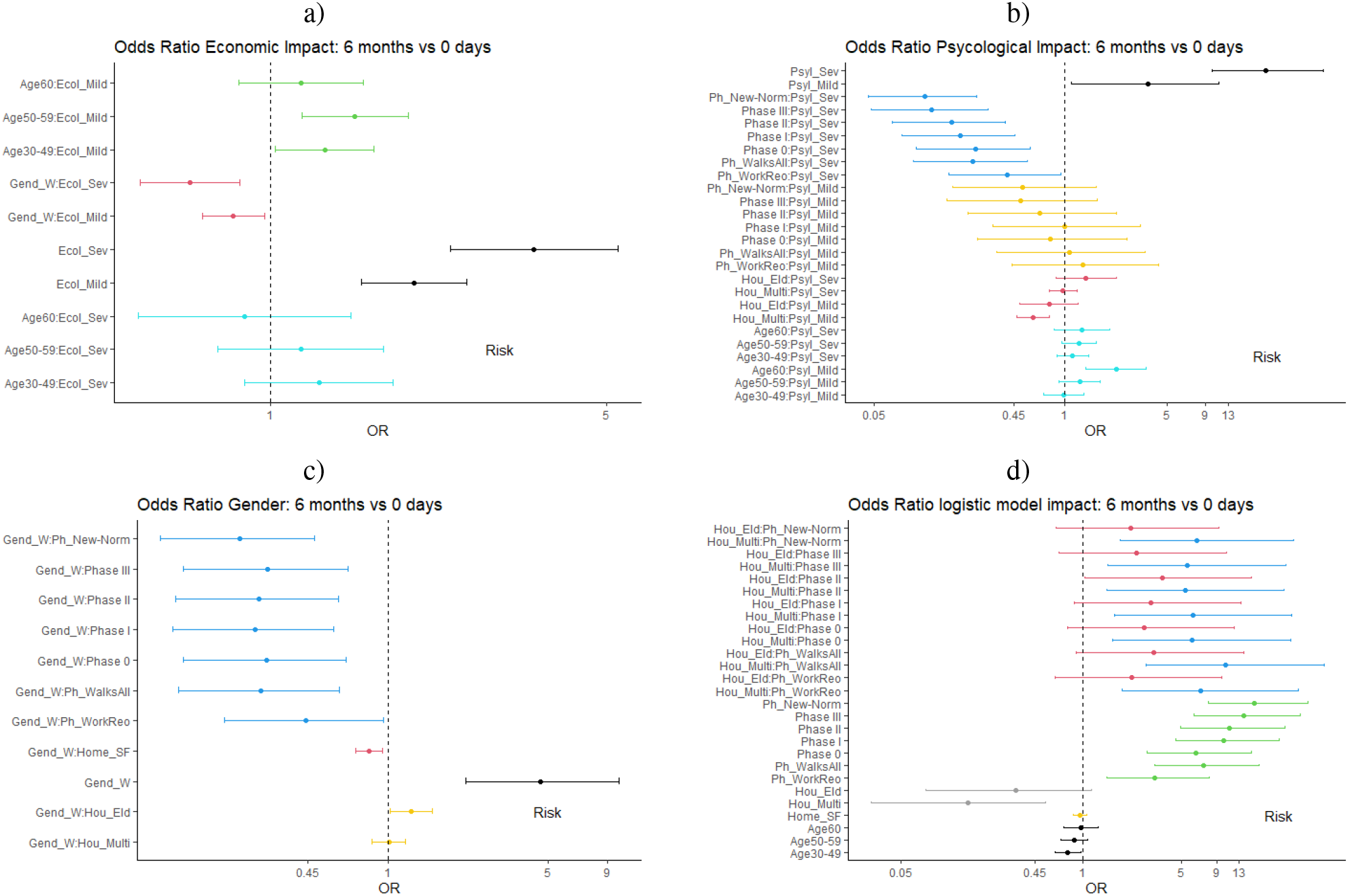
Odds Ratios for all the variables with interactions in the logistic regression model.

### Economic Impact

Figure 1a) shows the OR for the *EcoI* variable taking as basal category *None*. Note how the OR is larger than one for all values of the *EcoI* variable, meaning that this variable is a risk factor. Moreover, the OR for *EcoI = Severe* (3.53) is larger than for *EcoI = Mild* (1.99) (in black color): those who report having had severe economic impact are more likely to report that they are not willing to be confined when compared to those with mild economic impact.

In addition, the OR of the interaction *Age:EcoI* (cyan lines for Severe and green for Mild) is larger than 1 for those aged 30-59 years old. Thus, people in that age group are more likely to report that they are not willing to be confined than those aged 18-29 years old (basal category). In the case of respondents aged 60+ years old, the OR is greater than one only when the economic impact is mild.

In terms of the *Gender:EcoI* interaction, the OR are less than 1 for both values of *EcoI* (red line). Hence, women with any kind of economic impact are less likely to report not willing to be in confinement any longer when compared to men (basal category).

### Pyschological Impact

Figure 1b) depicts the OR for the *PsyI* variable taking as basal category *None*. As shown in the Figure, those who report *(Severe or Mild)* psychological impact have higher risk to report an unwillingness to be confined than those without any psychological impact. Moreover, the larger the psychological impact, the higher the risk (black lines). In terms of the interaction *Age:PsyI*, all the age groups show a similar level of risk (cyan lines), except for those aged 60+ years old with mild psychological impact who seem to be at significantly higher risk of reporting that they would not be able to be in confinement any longer (OR = 2.23). We observe a difference in the risk depending on the age of the members of the *Household*: households composed of elderly (60+) are significantly more at risk to report an unwillingness to be in confinement when compared to the rest (red and cyan lines) Concerning the interaction between *Phase:PsyI*, it seems that time has an attenuating effect on the risk as there is a consistent decrease of the risk over time. These results occur for *PsyI = Severe* (blue lines) and *PsyI = Mild* (yellow lines) but in this case to a lesser extent. This finding is surprising, as one would expect people to be at higher risk of not tolerating confinements the longer the pandemic lasts.

### Gender

In Figure 1c) the OR for the *Gender*_*W*_ variable are shown, taking *Men* as the basal category. As depicted in the Figure, women tend to be at significantly higher risk (OR = 4.60) than men to report that they are not able/willing to be in confinement any longer (black line). The value of this OR is significantly larger than the OR found for the *EcoI* and *PsyI* variables. This risk is increased for women who live in households composed of elders *Household*_*Elder*_ (yellow lines). In terms of the interaction *Phase:Gender*_*W*_, all the OR are similar and lower than 1 (blue lines), meaning that as time goes by, the risk for women to report that they are not able to be in confinement is lower than during the reference phase (*Workplace closure*).

### Rest of Variables

Figure 1d) depicts the OR of the rest of variables and their interactions. As seen in the Figure, the OR of *Phase* as a main factor is larger than one for all its values when compared to its basal value (*Workplace closure*) and increases over time from 3.3 during the *Work reopening* phase to 16.7 during the *New normality* phase. In other words, the risk of reporting an unwillingness to stay in confinement increases significantly as time goes by.

Regarding the type of *Household*, multi-generational households are at lower risk of reporting that they are not willing to be confined any longer than households with composed of elders (grey lines). However, in both cases the OR < 1, so they are attenuating factors when compared to the basal category (*Household = Young*).

We do not observe a significant impact of *Age* as a main factor on the risk to report being unwilling to be confined any longer. Nonetheless, as previously reported, age can be a risk factor in its interaction with other factors, such as *Economic* and *Psychological Impact*.

Combining all the OR reported above, we conclude that men living in multi-generational homes, with neither psychological nor economic impact due to the pandemic, are the least likely group to report an unwillingness to be confined. Conversely, women aged 30+ years old with severe economic and psychological impact and living in households with young children are at the highest risk to report an unwillingness to be confined.

### Classification Performance

Given that we have a quantitative model of the unwillingness to be confined, we test the model as a binary classifier to automatically infer such unwillingness from our independent variables. We randomly split the data in a training (80% of the data) and testing (20% of the data) sets. Note that we group the data on a weekly basis to have enough samples of survey answers across the entire time period of study.

On this binary classification task, the logistic regression model has a sensibility of 72.5% *±* 0.73 and a specificity of 63.6% *±* 0.75 with 100 random permutation data for training. A random model would have a sensibility of 50.0% *±* 0.004 and a specificity of 49.99% *±* 0.005. We experimented with other state-of-the-art machine learning methods. Gradient Boosted Trees (GBTs) trained following the same sampling methodology obtained a sensibility of 58.0% *±* 0.01 and a specificity of 60.7% *±* 0.01. The three most important variables identified by the GBTs were *Psychological impact, Phase* and *Economic impact*.

### Automatic Pattern Extraction

In addition to the logistic regression modeling, we have developed an automatic pattern extraction algorithm described in the Materials and Methods Section. Using this model, we identify the most influential variables and discover the most recurrent patterns among those who report an unwillingness to be confined.

### Most Influential Variables

Figure 2 depicts the variable importance obtained with our pattern extraction methodology across the different phases of the pandemic. As displayed in the Figure, there is a significant change over time of the importance of two variables: the *Psychological (PsyI)* and *Economic Impact (EcoI)* of the pandemic on the survey respondents.

**Figure 2.**
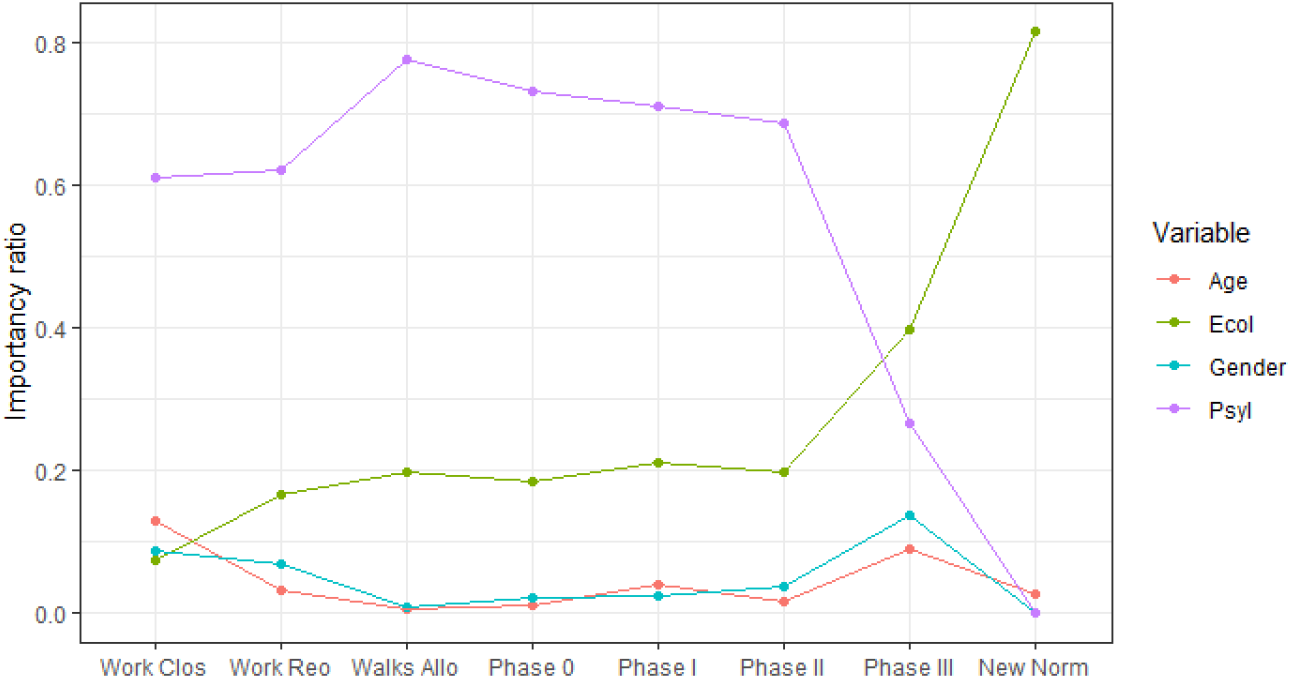
Evolution of the variable importance over time, according to the pattern extraction model.

Initially, *PsyI* is the most important variable, increasing its importance up to the *Walks allowed* phase, when its importance begins to decline. Conversely, the *EcoI* variable is not important at the beginning of the pandemic. Up until the end of *Phase II*, the importance of this variable increases smoothly or remains constant over time. However, with the start of *Phase III* and the return of the new normality, the importance of the economic impact to determine the unwillingness to be confined becomes evident.

The curves corresponding to the importance of the psychological and economic impact variables intersect between *Phase II* and *Phase III*. The rest of the variables exhibit a fairly stable behaviour over time, with mild fluctuations in their importance, affecting the target variable almost uniformly throughout the different phases of confinement.

Given this dependency of the target variable on the Phase of the pandemic, we discuss next our findings when performing an analysis in each of the 8 Phases of study (see Figure 5).

### Analyses per Phase of the Pandemic

#### Logistic Regression

In Figure 3 we show the OR of the main effects and their temporal evolution over time in each of the phases of the pandemic. Note that for each *Phase* we build a different logistic model with the main variables but without interactions given that we do not have enough data per Phase to identify meaningful interactions.

**Figure 3.**
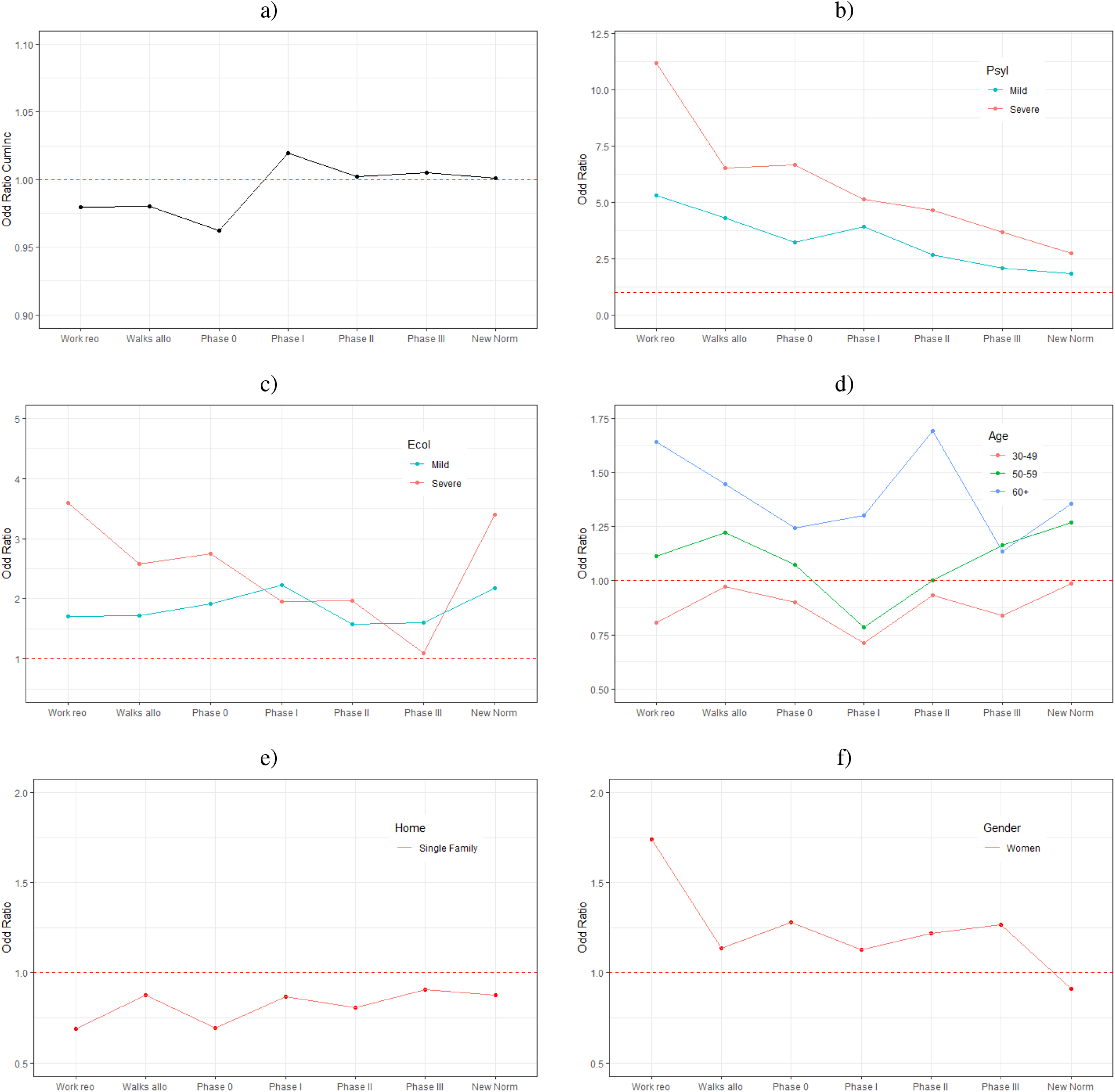
Odds Ratios for the main variables of the models per phase. a) CumInc b) Psychological Impact, c) Economic Impact, d) Age, E) Type of Home and F) Gender. Each point on the X axis corresponds to the OR of the logistic model for that particular phase.

Figure 3 a) shows the Odds Ratio of the 14-day cumulative incidence over time. The OR increases over time, which might be indicative of the so called *pandemic fatigue*. In the early phases of the pandemic, the OR is less than one, meaning that the larger the 14-day cumulative incidence, the lower the probability for respondents to report an unwillingness to be in confinement. Conversely, from *Phase II* onward, respondents’ seem to be indifferent (OR close to 1) to changes in the cumulative incidence. Figure 3 b) a depicts the OR of the *Psychological Impact (PsyI)* variable taking as basal category *No impact*. As seen in the Figure, the OR are always larger than 1 and decrease over the time. Hence, the role that the psychological impact variable plays in increasing the probability of reporting not being able to be confined any longer decreases as the months go by. This finding is consistent with the previously reported variable importance analysis (see Figure 2).

In the case of the *Economic Impact (EcoI)* variable (Figure 3 c)), the OR are also larger than 1 in all the phases. However, the evolution over time is different than that of the *PsyI* variable. We observe a significant increase in the OR after the *Phase III*, particularly for those reporting severe economic impact. This result is also consistent with the variable importance analysis previously described. Moreover, it makes intuitive sense: as the economic situation of people worsens due to the pandemic, their probability of reporting that they are not willing to be in confinement any more increases.

The changes over time in the OR for the *Age* variable are depicted in Figure 3 d). Those aged 60+ are at significantly larger risk in the first phases of the pandemic to say that they “can’t anymore” when compared to the rest of age groups. Once the *New normal* phase started at the end of June of 2020, the risk of those aged 50-59 and those aged 60+ becomes similar. Interestingly, the 30-49 age group is the only age group with an OR < 1 during the entire period.

Those living in single family homes are at lower risk to report that they cannot be in confinement any longer when compared to those living in an apartment (Figure 3 e)). This finding makes intuitive sense as single family homes are typically more spacious than apartments in Spain.

Finally, *women* are at larger risk than men to report that they cannot endure the confinement any longer throughout the entire period until reaching the *New normal* phase, when the OR for women becomes smaller than 1 Figure 3 e).

### Automatic Pattern Detection

Given the changes of the most relevant variables over time (see Figures 2,4), we apply our automatic pattern extraction algorithm to the data in each of the phases. We extract the patterns that model the unwillingness to be confined from the two most influential variables: economic and psychological impact, together with gender and age. Figure 2 shows a graph comparison of the weights of the most important variables as per our pattern detection method applied in each of the phases of confinement. As seen in the Figure, each confinement phase is modeled differently by these explanatory variables. Thus, the patterns identified in each phase change as the variables become more or less important.

**Figure 4.**
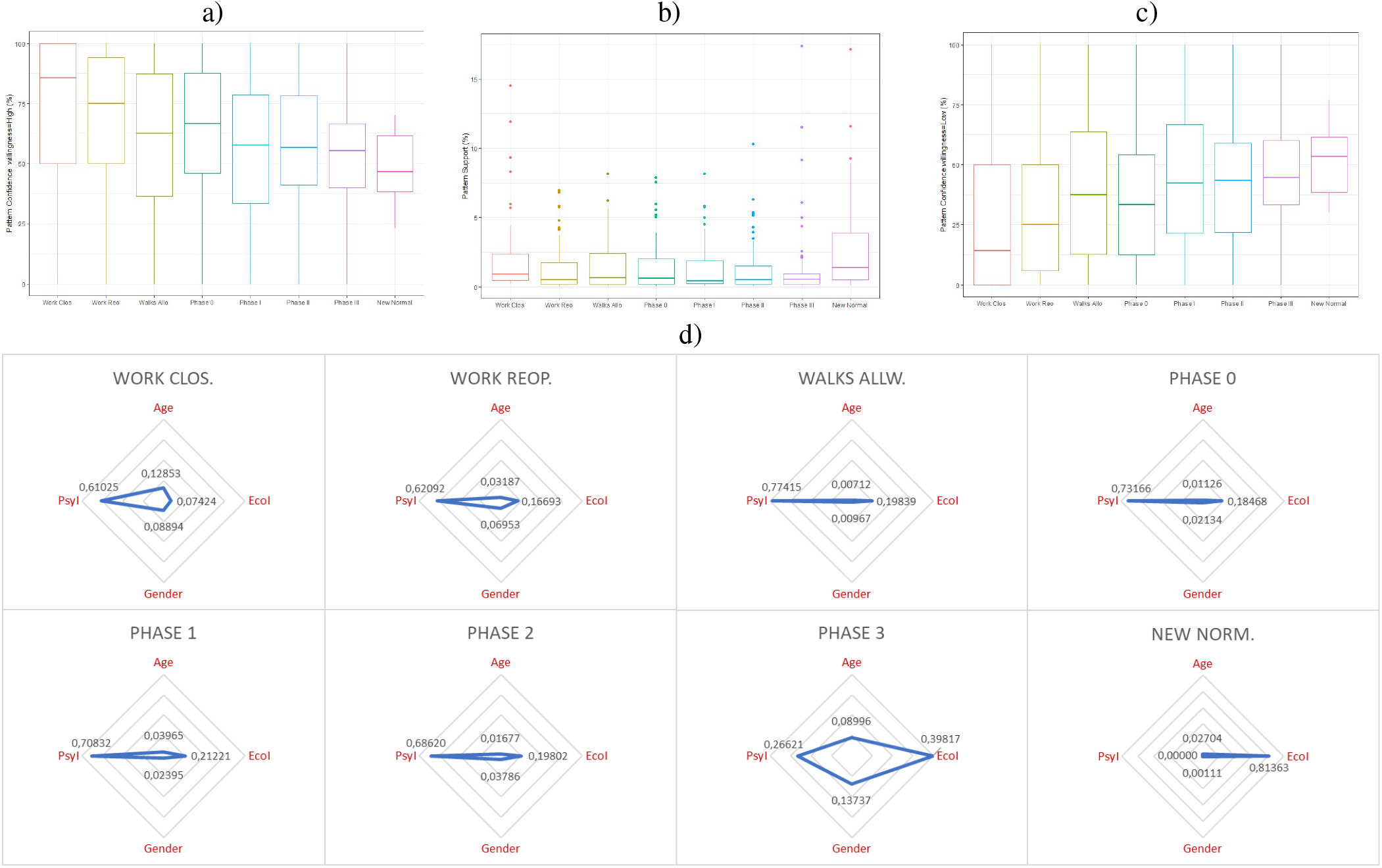
a) Confidence statistics of the patterns in each phase for the willingness to be confined for 6 months or longer. b) Support statistics of the identified patterns in each phase. c) Confidence statistics of the patterns in each phase for the willingness to be confined for 0 days. d) Weights of the most important variables according to our pattern detection method per confinement Phase.

As shown in Figure 4, age and gender have relatively important weights in the first stage of confinement but then practically disappear until *Phase III* to disappear again in the *New normal* phase.

The intermediate phases of confinement (from *Walks allowed* to *Phase II*) can be mainly modeled using the *Psychological Impact* variable. In *Phase III* (the last stage prior to the *New normal*), the unwillingness to be confined is affected by the two socio-demographic variables (gender and age), by psychological impact and especially by economic impact, which is triggered in this phase. In the *New normal* phase, the unwillingness to be confined is mainly dependent on economic impact.

Next, we summarize the most relevant patterns identified in each of the phases.

### 1. Workplace closure

In this phase, those over 50 years of age, regardless of their gender and economic impact, as long as they report no psychological impact (26.4% of the sample) always opt (100% of the time) for a willingness to be confined for 6 months or longer.

While we did not obtain any significant interaction between gender and psychological impact in our logistic regression modeling, we did identify significant gender differences in the 30 - 49 years old age range: among those who do not report any economic impact but do report severe psychological impact (close to 10% of the sample): 82.6% of men vs 64.7% of women would be willing to be in confinement for 6 months or longer.

This finding illustrates the complementary nature of our modeling approaches. The pattern discovery method identifies patterns that involve three or more variables, which would be very difficult to achieve via our logistic regression methodology.

In this initial stage of confinement, when the psychological and/or economic impact might not yet evident, the willingness to be confined is at its highest levels. However, 2.6% of the sample corresponds to women between 50 and 59 years old who report severe psychological impact. Of these, 60.0% report an unwillingness to be in confinement, even in this very early stage of the pandemic.

### 2. Workplace reopening

A large percentage (close to 94% on average) of men and women between 30 and 60 years old, who report neither economic nor psychological impact (18.4% of the sample) are willing to be confined for 6 months or longer. When this group reports mild psychological impact (13.1% of the sample), we observe gender differences, with 92.5% of men vs 85.4% of women willing to remain in confinement for 6 months or longer.

As expected, the reopening of workplaces has hardly any impact on people aged 60+ (mostly of non-working age) without economic or psychological impacts: 94.1% on average of them report a willingness to be confined for 6 or more months, irrespective of their gender. This group represents 11.6% of the sample.

At this stage, those who report high levels of acceptance towards confinement still clearly outweigh those unwilling to be confined. However, once again, we identify clear gender differences in the patterns, with women being more likely to report that they cannot be in confinement any longer when compared to men. In particular, we identify two distinct patterns: first, 43.6% of women between 50 and 59 years old who report severe psychological impact and without economic impact (3.7% of the sample) report minimal acceptance towards confinement; second, 56.5% of women between 30 and 49 years old, who in addition to reporting severe psychological impact also claim to have medium economic impact (2.7% of the sample) also respond that they would not be able to continue in confinement.

### 3. Walks allowed

During this phase, the participants’ opinions regarding their willingness to be confined starts to balance: the percentage of those willing to be confined for 6 months or longer decreases whereas those who are not willing to be confined increase.

The first identified pattern corresponds to people between 30 and 49 years old who report having no economic impact but severe psychological impact (13.0% of the sample). Among these (men and women alike), 54.8% on average report a willingness to be confined for 6 months or longer.

At this stage, there are several groups (both men and women) that begin to opt mainly for an unwillingness to be confined. The pattern with the largest support reveals that 75.0% of women between 30 and 49 years old with medium economic impact and a severe psychological impact (3.7% of the sample) report an unwillingness to be confined.

### 4. Phase 0

In the first phase of the re-opening in Spain, more people report being able to continue in confinement for 6 months or longer than those reporting that they cannot stand it any longer, with the exception of two groups: women between 30 and 49 with severe psychological impact and mild economic impact (3.49% of the sample) and women over 60, with severe psychological impact but no economic impact (6.0% of sample). In these two groups, only 46.9% and 45.2% respectively report a willingness to be confined for 6 months or longer.

While the willingness to be in confinement for 6 months or longer also decreases among men, still remains the most popular option for them. This finding is aligned with our OR analysis shown in Figure 1 c).

Half of those aged 60+ (both men and women) who do not declare to have any economic impact but report severe psychological impact (9.0% of the sample) report not being able to remain in confinement any longer. The burden of the pandemic starts to become evident.

### 5. Phases I and II

Both phases show similar patterns where the unwillingness to be confined is prevalent between individuals who report severe psychological impact with some economic impact. These levels of unwillingness are higher when, in addition to the severe psychological impact, participants also report some type of economic impact.

In Phase I, the opinion of some groups is polarized clearly towards an unwillingness to be confined: 67.2% of women between 30 and 59 years old who report medium economic impact and severe psychological impact (over 7.4% of the sample) and 86.9% of women aged 60+ who, having no economic impact declare severe psychological impact (5.0% of the sample) respond that they cannot stand the confinement any longer. Again, the most significant patterns related to those who are unwilling to continue in confinement are found amongst women.

### 6. Phase III

In the last phase of the re-opening and before the imminent start of the *New normal* phase, we observe an increase among those who report being able to remain in confinement for 6 months or longer. As seen in our feature importance analysis, the psychological impact becomes less relevant than economic impact and demographic factors. Hence, psychological impact does now appear in most of the patterns.

In general terms, the majority opinion is once again towards a willingness to be confined for 6 months or longer. The group that reports the highest levels of unwillingness to be in confinement (47.3% of the time) are people (men and women) between 30 and 49 years old who do not report psychological impact but report economic impact (7.6% of the sample).

### 7. New Normal

The willingness to be confined shows a balanced distribution between those who affirm that they are not able to stand it any longer and those who would be willing to be confined for 6 months or longer: 65.2% of people between 30 and 50 years old (without distinction of gender) who report neither economic nor psychological impact (28.8°% of the sample) would accept long-term confinement, compared to 34.8% who would not stand confinement any longer.

During this period (between the end of June 2020 and September 2020), the economic impact is the most influential factor to determine the willingness to be confined, when compared to gender, age and psychological impact. We highlight two patterns that strongly support (in more than 67.7% of the cases) an unwillingness to be confined: women aged between 30 and 49 years old who do not report psychological impact, but report severe economic impact (3.4% of the sample), and men aged between 50 and 59 years old, who do not report psychological impact either, but who report moderate economic impact (2.7% of the sample). Both groups are of working age and with some level of economic impact. They clearly opt for an unwillingness to be confined.

## Discussion

We have described two different analytical approaches applied to the same data. The obtained results are consistent yet also complementary with each other. The logistic regression modeling revealed the main factors that impact the unwillingness to be confined and enables us to build a predictive model of such an unwillingness. The qualitative methodology automatically identified relevant patterns that characterize the groups of respondents who exhibit an unwillingness to be in confinement.

From these analyses, we draw several key findings related to the interplay between demographic factors, the economic and psychological impact of the pandemic on people’s lives and the passage of time.

### 1. Pandemic fatigue is evident

Our sample spans over a 5-month period which includes both the severe lockdown imposed by the Spanish government between March 2020 and June 2020 and more than two months of the *New normal* phase. As the weeks go by, we observe a clear decrease of those who report a willingness to be confined for 6 months or longer from 85.8% to 55.4% of the sample. We corroborate this finding in our OR analysis, where the risk to report an unwillingness to be in confinement any longer significantly increases with the *Phases* of the pandemic (see Figure 1 d)). The most important variables to determine the unwillingness to be confined change as the months go by: psychological impact starts as the most important variable whereas economic impact becomes the most important variable once the *New normal* phase starts. Moreover, until *Phase 0* of the pandemic, the larger the 14-day cumulative incidence of COVID-19 cases, the lower the risk of reporting an unwillingness to be confined. However, as the months went by, the impact of the cumulative incidence on the unwillingness to be in confinement disappeared, starting in *Phase II* of the pandemic.

Since the World Health Organisation published its report on pandemic fatigue^34^, several authors have investigated the topic. Reicher and Drury propose that the non-adherence to the confinement measures is not due to pandemic fatigue, but rather due to structural weaknesses in our society, such us the inability to effectively self-isolate^35^. Conversely, other authors have written about the psychological, physical and economic impact of the pandemic on people, which has led to the emergence of the pandemic fatigue in different countries in the world^36–41^.

Given that the SARS-CoV-2 pandemic is expected to last for many more months, new confinements might be needed to control the transmission rate of the disease. However, the population is in a very different mental and economic state than they were at the beginning of the pandemic. Hence, policy makers should be sensitive to this situation in order to maximize the chances that future confinement measures will be both accepted and complied with by the population.

### 2. Gender and age matter

We identify very significant gender differences in the reported unwillingness to be confined with women being more likely to be unwilling throughout all the phases of the pandemic. According to the OR analysis, age does not seem to play a significant role in increasing the risk to report an unwillingness to be confined. However, our qualitative pattern discovery method identifies several patterns involving individuals (and especially women) of working age with some level psychological and/or economic impact and a clear unwillingness to remain in confinement any longer.

Previous work has also identified significant gender differences, with women being generally more compliant with individual protection and confinement measures^42,43^ than men. Galasso et al. report that women are more likely to perceive the pandemic as a very serious health problem and to agree and to comply with restraining measures from the analysis of survey data in eight countries conducted in March-April 2020^44^. Haischer et al.^45^ studied facial mask wearing behavior in a sample of 9,935 shoppers and identified that women were 1.5 times more likely to wear a mask than men. The fact that women in our study are at larger risk of not being willing to be in confinement when compared to men might be explained by the fact that women also are more likely to report psychological impact. According to our data, 18.0% of men vs 20.8% of women and 25.0% of men vs 30.6% of women (*χ*^2^ = 143.45, p-value < 2.2e-16) report mild and severe psychological impact due to the pandemic, respectively. Previous research has also studied the differential psychological impact of the pandemic on women^46–48^.

This gender and age-based differences are important to policy makers, as they reveal the need for gender and age-dependent public policies that would address the needs of different gender and age groups.

### 3. From psychological to economic impact

Psychological impact played a very important role in the early stages of the pandemic (up to *Phase III*): it is the main variable that determines the unwillingness to be confined, as reflected in our OR and feature importance analyses. However, as the months went by, psychological impact became less relevant whereas the economic impact of the pandemic became the main driver to determine an unwillingness to be confined. In the *New normal* phase, those with severe economic impact are significantly more likely to report that they cannot stand the confinement any longer when compared to those without any reported economic impact.

Other authors have studied the psychological impact of the COVID-19 pandemic on different populations, including students ^49^, healthcare workers ^50^, and the general population^51–53^. The economic impact of pandemics throughout history and for the COVID-19 pandemic has been analysed by several authors, including Ceylan et al.^54^ who conclude that unemployment with its income effects will be the most prominent economic consequence of the pandemic. However, we have not found any previous work that has studied the evolution over time of the role played by both the psychological and economic impact on the willingness to be in confinement.

This finding points out the importance of deploying both psychological and financial support measures as people’s livelihoods are severely impacted by over one year of the coronavirus pandemic.

## Conclusions

Population confinements have been one of the most widely adopted non-pharmaceutical interventions (NPIs) implemented by governments across the globe to help contain the spread of the SARS-CoV-2 virus. While confinement measures have been proven to be effective to reduce the number of infections, they entail significant economic and social costs. Thus, understanding the factors that determine the willingness of individuals to be confined during a pandemic is of paramount importance, particularly, to policy and decision-makers.

In this paper, we have studied such factors by means of a large-scale, online population survey deployed in Spain, one of the most affected countries in Europe by the pandemic. We consider socio-demographic, economic and psychological factors, together with reported 14-day cumulative incidence per 100,000 inhabitants. We analyze 109,515 answers provided during a 5-month period (between April 3rd, 2020 and September 11th, 2020) to shed light on the impact of the passage of time.

We apply both quantitative (logistic regression) and qualitative (pattern identification) methodologies to determine the role that the factors play in determining the unwillingness to be confined and their evolution over time. We find evidence of *pandemic fatigue*: the unwillingness of people to be confined increases as the months go by. We identify significant gender and age differences, placing women –and particularly working age women– at a higher risk than men to be unwilling to be confined. Finally, we uncover the importance of psychological impact at the early stages of the pandemic to be replaced by economic impact once the *New normal* was declared at the end of June of 2020.

Our results point to the need to design age and gender specific public policies, to implement economic programs to support the many citizens whose livelihoods have been severely impacted by the pandemic and to address the evident pandemic fatigue as the success of potential future confinements will depend on the population’s willingness to comply with them.

Collecting a large sample of survey data does not come without limitations. First, respondents have to be adults (at least 18 years old). Second, there might be a difference between the reported willingness to be in confinement and the actual compliance with the confinement measures. Third, there is a self-selection and sampling bias as the survey is filled out by volunteers who have learned about the survey via social media, WhatsApp, newspapers articles or Facebook ads, and who need access to a computing device (smartphone, tablet, PC) with internet connection. We mitigate this bias by weighing the raw data and by deploying gender-balanced Facebook advertisement campaigns.

## Material and Methods

### The COVID19Impact Survey

We report the results of analyzing a subset of the answers to the COVID19ImpactSurvey, a very large, anonymous, online citizen survey about COVID-19^33^. Launched on March 28th in Spain, the survey has since collected over 600,000 anonymous answers mainly from 4 countries (Spain, Italy, Brazil and Germany). From this very large sample, we analyze the data from Spain for the time period between April, 3rd 2020 and September, 11th 2020.

The COVID19ImpactSurvey was launched in collaboration with the Valencian Government in Spain and has been regularly used to assist in their decision-making^33^. The survey has changed over time to adapt to each of the phases of the pandemic. During the period of our study, the survey consisted of 25 questions designed to collect information about the participants’ demographic and household situation (Q1-Q7); their social contact behaviour (Q8-Q12); their support for the government measures deployed to contain the spread of coronavirus (Q13) and their willingness to continue in confinement (Q14); the economic and labor impact of the pandemic in their lives (Q15-Q19); their health risk factors (Q20), their ability to self-isolate if needed (Q21), the presence of coronavirus-related symptoms (Q22), the duration of the symptoms (if any) (Q23), whether they have been tested for coronavirus (Q24); and the psychological/emotional impact of the pandemic in their home (Q25).

We refer the interested reader to^33^ for a detailed description of the survey questions, the weighing methodology applied to reduce biases in the data and the results of analyzing the first week of answers. Note that question Q25 related to the psychological impact of the pandemic was added on April 3rd and thus it is not included in the analysis presented in^33^.

From the questions described above, we report the results of analyzing the subset of questions depicted in S.I. Table ??, namely Q1-Q6, Q14, Q15 and Q25.

Given that we have data for several months, we analyze the survey answers according to the different phases of confinement implemented in Spain between April and September of 2020. Figure 5 illustrates the chronology of each of the phases as per official data^1^, together with the daily number of confirmed COVID-19 cases. The shaded portion of the graph corresponds to the time period of analysis in this paper.

**Figure 5.**
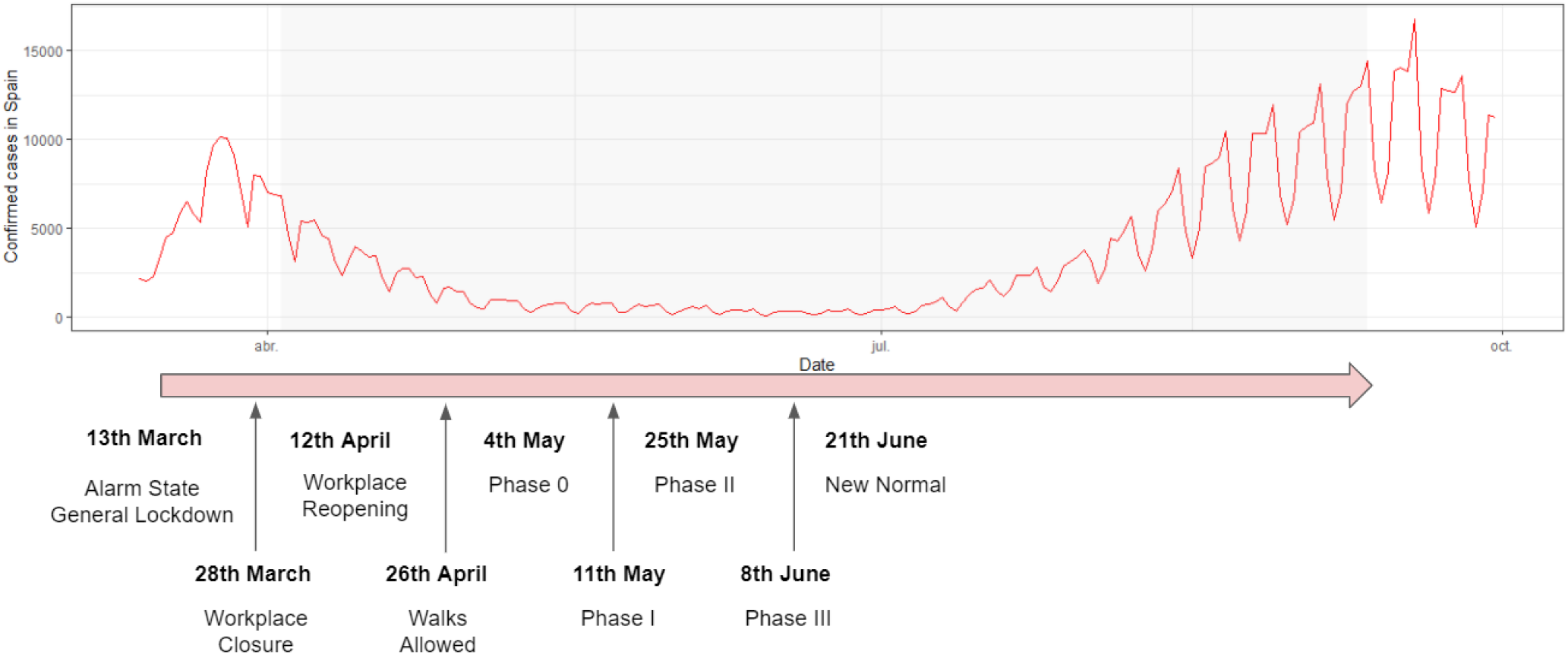
Confinement phases and daily number of confirmed COVID-19 cases in Spain between March and September of 2020. The shaded part of the graph corresponds to the time period of analysis.

### Data Processing

Table 1 summarizes the independent and dependent variables used in our study. Our target or dependent variable is the willingness to be confined, captured by Q14. Our independent variables are the socio-demographic, economic and psychological impact measures, captured by questions Q1-Q6, Q15 and Q25, the confinement Phase as shown in Figure 5, and the 14-day Cumulative Incidence of SARS-CoV-2 per 100.000 inhabitants averaged over the 17 Autonomous Regions in Spain.

To minimize biases in the data, we weigh the raw answers by gender, age and province, such that the resulting distribution of answers matches the distributions reported by the official Spanish census. We apply the same methodology as described in^33^. Table 2 in the Supplementary Information depicts the age and gender distributions of the raw and weighted data, together with the officially reported census distributions.

In addition, we simplify several of the answers as follows:

1. Q4. We only consider two types of homes: apartments and single family homes. These two answers represent 94% of the raw answers.
2. Q15. We group the answers to the economic impact question into three levels as follows: (a) *None (0)*: No economic impact is reported; (b) *Mild (1)*: I lost my job” or “I lost part or all of my savings” or “My business is in danger of bankruptcy” or “My employer is at risk of bankruptcy”; (c) *Severe (2)*: “I can’t afford to buy food” and/or “I can’t pay my rent or mortgage anymore”.
3. Q25. We group the answers to the psychological impact question into three levels: (a) *None (0)*: No psychological impact is reported; (b) *Mild (1)*: “Excessive use of technology by adults (tablet, phone, TV)” and/or “Excessive use of technology by children (tablet, phone, TV)”; (c) *Severe (2)*: “High levels of anxiety” and/or “High levels of stress” and/or “High levels of loneliness” and/or “High levels of sadness” and/or “Loud arguments or fights with other members of the home” and/or “Excessive consumption of alcohol” and/or “Excessive consumption of drugs (prescription or other)”.
4. Q14. We divide the target variable, *i*.*e*., the willingness to be in confinement into two categories, as per the answers to question Q14: (a) *Unwilling (coded as 0)*: Willingness to be confined for 0 days, the person cannot stand it anymore; (b) *Long-term (coded as 1)*: Willingness to be confined for at least 6 months.

### Logistic Regression Models

We built an explainable logistic regression model^55^ to predict the unwillingness to be confined (binary variable) using the variables described in Table 2 –namely, *Phase, Age, Gender, Home, Household, Economic Impact (EcoI), Psychological Impact (PsyI) and 14-day COVID-19 Cumulative Incidence (CumInc)*– as our independent variables with plausible interactions among them.

Note that in this regression model, we reversed the coding of the target variable, *i*.*e*. we coded with a value of 1 the willingness to be confined for zero days (*i*.*e*. the unwillingness to be confined), and with a value of 0 the willingness to be confined for at least 6 months. We used this reverse coding, such that our target variable represents the *risk* for respondents not to accept a confinement, thus being consistent with the risk analyses that are typically done in logistic regression modeling via an Odds Ratio (OR) methodology.

First, we built a complete model that included all the variables and interactions between them. Next, we performed feature selection by means of a bidirectional step-wise variable selection process, such that at each step we added or removed a main variable or an interaction according to the Akaike Information Criterion (AIc)^56^. Finally, we computed the p-value of all the remaining variables. If the p-values were not lower than 0.05 we performed a likelihood ratio test^57^ between the model with the variable included and the model without the variable to determine whether to include such a variable in the final model or not. We also used this model to automatically predict the target variable. Moreover, we carried out an Odds Ratio analysis to shed light on the role of each of the variables to determine the opposition to confinement, as described in the Results Section.

### Automatic Pattern Extraction

In addition to the logistic regression analysis, we performed a qualitative study to automatically identify meaningful patterns that would differentiate the behaviours, impact and characteristics of those not willing to be confined vs those willing to be confined for a long time period.

We followed a two-step methodology to automatically extract the patterns from the survey answers:

1. *Step 1: Selection of the most influential variables*. We identified the most influential variables with respect to our target variable (*i*.*e*., the willingness to be confined). Note that not all the variables in our study might be relevant when modeling people’s willingness to be confined. In this variable selection step, we used the Entropy-based Filters algorithm from the FSelectorRcpp library in R, which is based on the information gain, gain ratio and symmetrical uncertainty metrics.^58^
2. *Step 2: Automatic pattern generation*, via a modified version of the RBS algorithm^59^, a classifier based on the ID3 family^60^. The RBS algorithm is an iterative method that, without building the full structure of a tree, automatically identifies a set of rules (patterns) from a data set of discrete variables. The subsequent ordering and filtering of the patterns depends on the *significance* of the rule, which is a metric based on the classic concepts of support and confidence of the classification rules, but defined by intervals. We adapted the RBS algorithm to be used with re-weighted input samples rather than absolute supports, as it is usually done.

As a result of this two-step process, we obtained the set of rules or patterns that best model our target binary variable (willingness or acceptance to be confined). Each of the automatically identified patterns or classification rules has the structure shown in Figure 6. The Results Section describes both the most influential variables and the patterns identified by our qualitative analysis.

**Figure 6.**
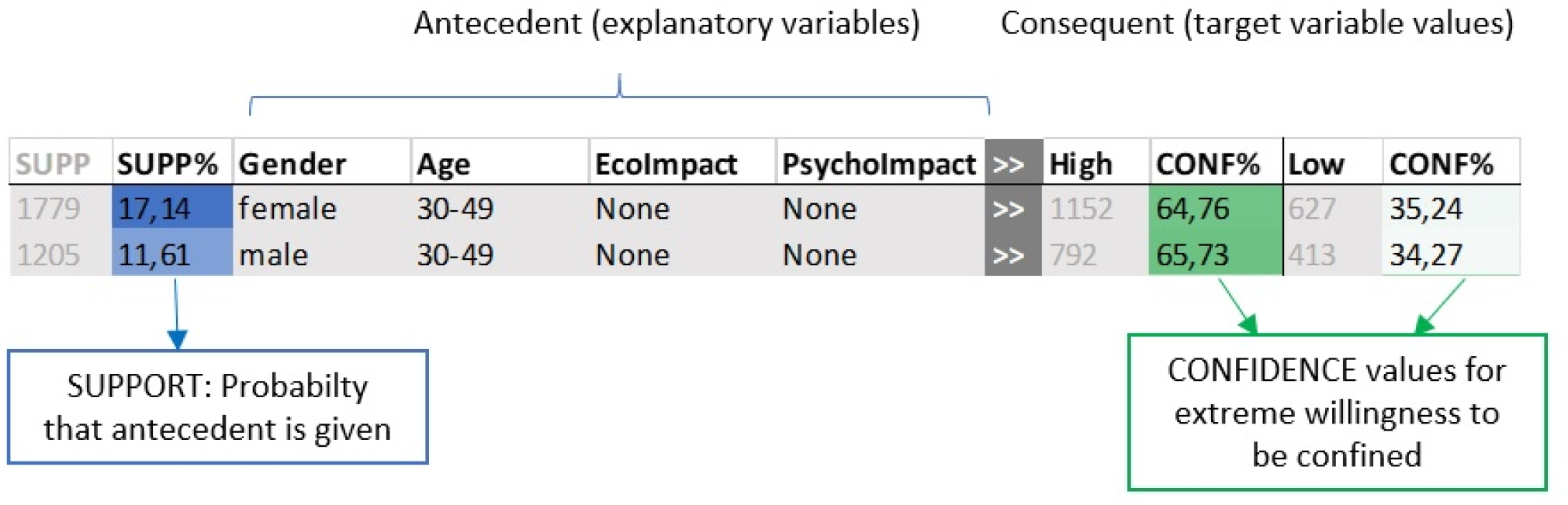
Example of two of the patterns identified by our pattern extraction method.

## Data Availability

The data will be made available upon publication in a peer-reviewed journal

https://ellisalicante.org/en/covid19impactsurvey

## Data availability

The survey answers analysed in this paper will be made available together with the manuscript.

## Code

The code used to analyse the data will be placed in a publicly available github server.

## Acknowledgements

This work is partially supported by grants FONDOS SUPERA COVID-19 Santander-CRUE, 2020–2021, Fundación BBVA a equipos de investigación científica SARS-CoV-2 COVID-19, IA4COVID19 2020-2022 and the Valencian Government.

## Author contributions

M.M-G. Contributed to the study design, performed the data processing and logistic regression modeling.

A.R. Contributed to the study design, performed the qualitative modeling and automatic pattern extraction.

X.B. Contributed to the study design, performed the data processing and logistic regression modeling.

K.P. Contributed to the study design, performed the qualitative modeling and automatic pattern extraction.

K.R. Contributed to the design and implementation of the online survey and raw data processing.

N.O. Contributed to the design and implementation of the online survey, the study design, the coordination of all the tasks, data analysis and insights extraction.

All authors contributed to the drafting of the manuscript.

## Additional information

This submission includes a Supplementary Information file.

Correspondence and request for materials should be addressed to N. O.

## Footnotes

1 See, for example, https://www.rtve.es/noticias/20200612/guia-fases-desescalada/2013020.html

## References

1. Zheng, Q. et al. Hit-covid, a global database tracking public health interventions to covid-19. Sci. data 7, 1–8 (2020).

2. Desvars-Larrive, A. et al. A structured open dataset of government interventions in response to covid-19. Sci. data 7, 1–9 (2020).

3. López, L. & Rodó, X. The end of social confinement and covid-19 re-emergence risk. Nat. Hum. Behav. 4, 746–755 (2020).

4. Lai, S. et al. Effect of non-pharmaceutical interventions to contain covid-19 in china. Nature 585, 410–413 (2020).

5. Verity, R. et al. Estimates of the severity of coronavirus disease 2019: a model-based analysis. The Lancet infectious diseases 20, 669–677 (2020).

6. Candido, D. S. et al. Evolution and epidemic spread of sars-cov-2 in brazil. Science 369, 1255–1260 (2020).

7. Day, T., Gandon, S., Lion, S. & Otto, S. P. On the evolutionary epidemiology of sars-cov-2. Curr. Biol. 30, R849–R857 (2020).

8. Betsch, C. et al. Social and behavioral consequences of mask policies during the covid-19 pandemic. Proc. Natl. Acad. Sci. 117, 21851–21853 (2020).

9. Gollwitzer, M., Platzer, C., Zwarg, C. & Göritz, A. S. Public acceptance of covid-19 lockdown scenarios. Int. J. Psychol. (2020).

10. Wang, Y. et al. The impact of quarantine on mental health status among general population in china during the covid-19 pandemic. Mol. psychiatry 1–10 (2021).

11. Liu, W., Yue, X.-G. & Tchounwou, P. B. Response to the covid-19 epidemic: the chinese experience and implications for other countries (2020).

12. Abiad, A., Arao, R. M. & Dagli, S. The economic impact of the covid-19 outbreak on developing asia. (2020).

13. Fernandes, N. Economic effects of coronavirus outbreak (covid-19) on the world economy. Available at SSRN 3557504 (2020).

14. Ivanov, D. Predicting the impacts of epidemic outbreaks on global supply chains: A simulation-based analysis on the coronavirus outbreak (covid-19/sars-cov-2) case. Transp. Res. Part E: Logist. Transp. Rev. 136, 101922 (2020).

15. Chen, D.-G., Chen, X. & Chen, J. K. Reconstructing and forecasting the covid-19 epidemic in the united states using a 5-parameter logistic growth model. Glob. Heal. Res. Policy 5, 1–7 (2020).

16. Clemens, V. et al. Potential effects of” social” distancing measures and school lockdown on child and adolescent mental health. Eur. Child & Adolesc. Psychiatry (2020).

17. Jiao, W. Y. et al. Behavioral and emotional disorders in children during the covid-19 epidemic. The journal Pediatr. 221, 264 (2020).

18. Sadati, A. K., B Lankarani, M. H. & Bagheri Lankarani, K. Risk society, global vulnerability and fragile resilience; sociological view on the coronavirus outbreak (2020).

19. Mukhtar, S. Psychological health during the coronavirus disease 2019 pandemic outbreak. Int. J. Soc. Psychiatry0020764020925835 (2020).

20. Gao, J. et al. Mental health problems and social media exposure during covid-19 outbreak. Plos one 15, e0231924 (2020).

21. Hawryluck, L. et al. Sars control and psychological effects of quarantine, toronto, canada. Emerg. infectious diseases 10, 1206 (2004).

22. Fischer, R. et al. Rapid review and meta-meta-analysis of self-guided interventions to address anxiety, depression, and stress during covid-19 social distancing. Front. psychology 11 (2020).

23. Colombo, R. A., Wallace, M. & Taylor, R. An essential service decision model for aba providers during crisis. Behav. Analysis Pract. 1 (2020).

24. Seale, H. et al. Covid-19 is rapidly changing: Examining public perceptions and behaviors in response to this evolving pandemic. medRxiv (2020).

25. Yamada, Y. et al. Covidistress global survey dataset on psychological and behavioural consequences of the covid-19 outbreak. Sci. data 8, 1–23 (2021).

26. Bailon, C. et al. Covidaffect: Real-time monitoring of mood variations following the covid-19 outbreak. (2020).

27. Amat, F., Arenas, A., Falcó-Gimeno, A. & Muñoz, J. Pandemics meet democracy. experimental evidence from the covid-19 crisis in spain. (2020).

28. Tintori, A. et al. Adaptive behavioural coping strategies as reaction to covid-19 social distancing in italy. Eur. review for medical pharmacological sciences 24, 10860–10866 (2020).

29. Landi, G., Pakenham, K. I., Boccolini, G., Grandi, S. & Tossani, E. Health anxiety and mental health outcome during covid-19 lockdown in italy: the mediating and moderating roles of psychological flexibility. Front. psychology 11, 2195 (2020).

30. McBride, O. et al. Monitoring the psychological impact of the covid-19 pandemic in the general population: an overview of the context, design and conduct of the covid-19 psychological research consortium (c19prc) study. (2020).

31. Freeman, D. et al. Coronavirus conspiracy beliefs, mistrust, and compliance with government guidelines in england. Psychol. Medicine 1–13 (2020).

32. Chan, H. F., Skali, A., Savage, D. A., Stadelmann, D. & Torgler, B. Risk attitudes and human mobility during the covid-19 pandemic. Sci. reports 10, 1–13 (2020).

33. Oliver, N., Barber, X., Roomp, K. & Roomp, K. Assessing the impact of the covid-19 pandemic in spain: Large-scale, online, self-reported population survey. J Med Internet Res 22, e21319. 10.2196/21319 (2020).

34. Organisation, W. H. Pandemic fatigue. reinvigorating the public to prevent covid-19. Policy framework for supporting pandemic prevention management (2020). https://apps.who.int/iris/bitstream/handle/10665/335820/WHO-EURO-2020-1160-40906-55390-eng.pdf.

35. Reicher, S. & Drury, J. Pandemic fatigue? how adherence to covid-19 regulations has been misrepresented and why it matters. BMJ 372, 10.1136/bmj.n137 (2021). https://www.bmj.com/content/372/bmj.n137.full.pdf.

36. Zerbe, K. J. Pandemic fatigue: Facing the body’s inexorable demands in the time of covid-19. J. Am. Psychoanal. Assoc. 68, 475–478, 10.1177/0003065120938774 (2020). PMID: 32589057, https://doi.org/10.1177/0003065120938774.

37. Al-Tammemi, A. B., Tarhini, Z. & Akour, A. A swaying between successive pandemic waves and pandemic fatigue: Where does jordan stand? Annals Medicine Surg. 65, 102298, https://doi.org/10.1016/j.amsu.2021.102298 (2021).

38. MacIntyre, C. R. et al. Mask use, risk-mitigation behaviours and pandemic fatigue during the covid-19 pandemic in five cities in australia, the uk and usa: A cross-sectional survey. Int. J. Infect. Dis. 106, 199–207, https://doi.org/10.1016/j.ijid.2021.03.056 (2021).

39. Seiter, J. S. & Curran, T. Social-distancing fatigue during the covid-19 pandemic: a mediation analysis of cognitive flexibility, fatigue, depression, and adherence to cdc guidelines. Commun. Res. Reports 38, 68–78, 10.1080/08824096.2021.1880385 (2021). https://doi.org/10.1080/08824096.2021.1880385.

40. Ilesanmi, O. S., Bello, A. E. & Afolabi, A. A. Covid-19 pandemic response fatigue in africa: causes, consequences, and counter-measures. The Pan Afr. medical journal 37(Suppl 1), https://doi.org/10.11604/pamj.supp.2020.37.37.26742 (2020).

41. Petherick, A. et al. A worldwide assessment of covid-19 pandemic-policy fatigue. (2021).

42. Petersen, E. et al. Covid-19–we urgently need to start developing an exit strategy. Int. J. Infect. Dis. (2020).

43. Kimhi, S., Marciano, H., Eshel, Y. & Adini, B. Resilience and demographic characteristics predicting distress during the covid-19 crisis. Soc. Sci. Medicine 265, 113389, https://doi.org/10.1016/j.socscimed.2020.113389 (2020).

44. Galasso, V. et al. Gender differences in covid-19 attitudes and behavior: Panel evidence from eight countries. Proc. Natl. Acad. Sci. 117, 27285–27291 (2020).

45. Haischer, M. H. et al. Who is wearing a mask? gender-, age-, and location-related differences during the covid-19 pandemic. https://doi.org/10.1371/journal.pone.0240785 (2020).

46. The impact of the covid-19 pandemic on women’s mental health. Arch Womens Ment Heal. 23, https://doi.org/10.1007/s00737-020-01092-2 (2020).

47. Gender dimensions of the covid-19 pandemic. World Bank Policy Note (2020).

48. Rodríguez-Rey, R., Garrido-Hernansaiz, H. & Collado, S. Psychological impact and associated factors during the initial stage of the coronavirus (covid-19) pandemic among the general population in spain. Front. Psychol. 11, 1540, 10.3389/fpsyg.2020.01540 (2020).

49. The psychological impact of the covid-19 epidemic on college students in china. Psychiatry research 287, https://doi.org/10.1016/j.psychres.2020.112934 (2020).

50. Psychological impact of the covid-19 pandemic on health care workers in singapore. Ann Intern Med. 173, 317–320, doi:10.7326/M20-1083 (2020).

51. Brooks, S. K. et al. The psychological impact of quarantine and how to reduce it: rapid review of the evidence. The lancet 395, 912–920 (2020).

52. Covid-19 pandemic and psychological fatigue in turkey. Int. journal social psychiatry advance online publication, 10.1177/0020764020941889 (2020).

53. Terry, P. C., Parsons-Smith, R. L. & Terry, V. R. Mood responses associated with covid–19 restrictions. Front. Psychol. 11, 3090 (2020).

54. Ceylan, R. F., Ozkan, B. & Mulazimogullari, E. Historical evidence for economic effects of covid-19 (2020).

55. Bruin, J. newtest: command to compute new test @ONLINE (2011).

56. Akaike, H. Information theory and an extension of the maximum likelihood principle. In Selected papers of hirotugu akaike, 199–213 (Springer, 1998).

57. Hosmer, D. W., Hosmer, T., Le Cessie, S. & Lemeshow, S. A comparison of goodness-of-fit tests for the logistic regression model. Stat. medicine 16, 965–980 (1997).

58. Fayyad, U. & Irani, K. Multi-interval discretization of continuous-valued attributes for classification learning. (1993).

59. Almiñana, M., Escudero, L., Pérez-Martín, A., Rabasa, A. & Santamaría, L. A classification rule reduction algorithm based on significance domains. Top 22, 397–418 (2014).

60. Quinlan, J. Indroduction of decision trees machine learning. Boston (NL): Kluwer Acad. Publ. 1, 86–106 (1986).

