## Supplementary material for "Yet another lockdown? A large-scale study on people’s unwillingness to be confined during the first 5 months of the COVID-19 pandemic in Spain"

### Supplementary Information of the paper: Yet another lock-down? A large-scale study on people's unwillingness to be confined during the first 5 months of the COVID-19 pandemic in Spain

**Marina Martinez-Garcia**[1,†], **Alejandro Rabasa**[2,†], **Xavier Barber**[2,†], **Kristina Polotskaya**[2,†], **Kristof Roomp**[3], **and Nuria Oliver**[4*,†]

[1]Universitat Jaume I, Dept de matemàtiques, Castelló, 12071, Spain
[2]Universidad Miguel Hernandez, Centro de Investigación Operativa, Elche, 03202, Spain
[3]Microsoft Corporation, Washington, USA
[4]ELLIS Unit Alicante Foundation, Alicante, Spain
[†]these authors contributed equally to this work

#### ABSTRACT

This Supplementary Information file includes two tables with the questions analyzed in the survey and the distributions of answers per age and gender in the raw, weighted and census data.

| Question | Possible answers |
|---|---|
| **Demographic and Household information** | |
| Q1. What is your age range? | [18-20; 21-29; 30-39; 40-49; 50-59; 60-69; 70-79; 80+] |
| Q2. What is your gender? | [Male; female; another gender] |
| Q3. Postal code/Zip code | Numeric entry |
| Q4. Type of home | [Single family house; apartment/flat; shared apartment/flat; other shared accommodation; other] |
| Q5. Number of people in the home (including you) | [1; 2; 3; 4; 5+] |
| Q6. Age(s) of people in the home (excluding you, check all that apply) | [10 or less; 11-20; 21-29; 30-39; 40-49; 50-59; 60-69; 70-79; 80+] |
| **Willingness to be Confined (target variable)** | |
| Q14. If you are or had to be confined to not leaving your home, for how long could you stand it? | [0 days, I can't stand it anymore; 1 week; 2 weeks; 1 month; 2 months; 6 months] |
| **Economic Impact** | |
| Q15. What kind of economic impact has the coronavirus had on you? (check all that apply) | [I lost my job; I lost my savings; I can't pay my rent or mortgage anymore; I can't afford to buy food; I have lost most or all of my income; My business is in danger of bankruptcy; My employer is in danger of bankruptcy; None of the above] |
| **Psychological Impact** | |
| Q25. Have you noticed a significant increase in your home in any of the following areas that you consider damaging? (check all that apply) | [High level of anxiety; High level of stress; High level of loneliness; High level of sadness; Loud arguments or fights with other members of the home; Excessive consumption of alcohol; Excessive consumption of drugs (prescription or other); Excessive use of technology by adults (tablet, phone, TV); Excessive use of technology by children (tablet, phone, TV); I have not noticed a harmful increase in these areas; I prefer not to answer] |

**Table S1.** COVID19ImpactSurvey questions analysed in this study.

| | Unweighted | | Weighted | | Spanish census | |
|---|---|---|---|---|---|---|
| Gender | M | F | M | F | M | F |
| 18-20 | 63.5 % | 36.5 % | 50.7 % | 49.3 % | 48.99% | 51.01% |
| 21-29 | 67.6 % | 32.4 % | 51.2 % | 48.8 % | 48.99% | 51.01% |
| 30-39 | 65.3 % | 34.7 % | 44.4 % | 55.6 % | 49.89% | 50.11% |
| 40-49 | 63 % | 37 % | 46.1 % | 53.9 % | 49.45% | 50.55% |
| 50-59 | 63.7 % | 36.3 % | 49.2 % | 50.8 % | 50.44% | 49.56% |
| 60-69 | 58.6 % | 41.4 % | 49.4 % | 50.6 % | 51.94% | 48.06% |
| 70-79 | 51.3 % | 48.7 % | 57 % | 43 % | 54.67% | 45.33% |
| 80 + | 41.6 % | 58.4 % | 64.4 % | 35.6 % | 63.18% | 36.82% |
| Total | 62.4 % | 37.6 % | 50.7 % | 49.3 % | 50.9% | 49.01% |

**Table S2.** To minimise bias in the data, the raw survey answers have been weighted per gender and age in each of the 51 provinces of Spain, and by type of profession at a country level. The table shows the raw (left-most columns) and weighted (middle columns) distributions of answers per age and gender, compared to the officially reported census data (right-most columns).
